# Asymptomatic cases and limited transmission of SARS-CoV-2 in residents and healthcare workers in three Dutch nursing homes

**DOI:** 10.1101/2020.08.31.20185033

**Authors:** Laura W. van Buul, Judith H. van den Besselaar, Fleur M.H.P.H. Koene, Bianca M. Buurman, Cees M.P.M. Hertogh, the COVID-19 NH-STUDY GROUP

**Affiliations:** Department of Medicine for Older People, Amsterdam Public Health Research Institute, Amsterdam University Medical Center, Amsterdam, the Netherlands; Department of Internal Medicine, section of Geriatric Medicine, Amsterdam Public Health Research Institute, Amsterdam University Medical Center, Amsterdam, the Netherlands; Department of Medical Microbiology, Amsterdam University Medical Center, Amsterdam, the Netherlands; Department of Infectious Diseases, Public Health Laboratory, Public Health Service of Amsterdam, Amsterdam, the Netherlands; Public Health Service Hart voor Brabant, s’-Hertogenbosch, the Netherlands; Public Health Service of West-Brabant, Breda, the Netherlands; Tante Louise long-term care, Steenbergen, the Netherlands; Tante Louise long-term care, Bergen op Zoom, the Netherlands; Public Health Service of Rotterdam-Rijnmond, Rotterdam, the Netherlands; Argos Zorggroep long term care, Schiedam, The Netherlands; Department Infectious Disease Control, Public Health Service of Amsterdam, Amsterdam, the Netherland; Cordaan long-term care, Amsterdam, the Netherlands; Cordaan Geriatric Rehabiliation, Amsterdam, the Netherlands

## Abstract

**Purpose:** Many nursing homes worldwide have been hit by outbreaks of the new severe acute respiratory syndrome coronavirus 2 (SARS-CoV-2). We aimed to assess the contribution of a- and presymptomatic residents and healthcare workers in transmission of SARS-CoV-2 in three nursing homes.

**Methods:** Two serial point-prevalence surveys, 1 week apart, among residents and healthcare workers of three Dutch nursing homes with recent SARS-CoV-2 introduction. Nasopharyngeal and oropharyngeal testing for SARS-CoV-2, including reverse-transcriptase polymerase chain reaction (rRT-PCR) was presymptomatic or asymptomatic with standardized symptom assessment.

**Results:** In total, 297 residents and 542 healthcare workers participated in the study. At the first point-prevalence survey, 15 residents tested positive of which one was presymptomatic (Ct value>35) and three remained asymptomatic (Ct value of 23, 30 and 32). At the second point-prevalence survey one resident and one healthcare worker tested SARS-CoV-2 positive (Ct value >35 and 24, respectively) and both remained asymptomatic.

**Conclusion:** This study confirms a-and presymptomatic occurrence of Covid-19 among residents and health care workers. Ct values below 25 suggested that these cases have the potential to contribute to viral spread. However, very limited transmission impeded the ability to answer the research question. We describe factors that may contribute to the prevention of transmission and argue that the necessity of large-scale preemptive testing in nursing homes may be dependent of the local situation regarding prevalence of cases in the surrounding community and infection control opportunities.

**KEY SUMMARY POINTS:** *Aim:* To assess the contribution of a- and presymptomatic residents and healthcare workers in transmission of SARS-CoV-2 in three nursing homes by facility wide preemptive testing.

*Findings:* Occurrence of a-and presymptomatic residents and healthcare workers with Ct values below 25 was confirmed. However, evaluation of contribution to transmission of the virus was not possible because of limited positive cases in the follow-up.

*Message:* Necessity of large-scale preemptive testing for a- and presymptomatic SARS-CoV-2 cases in nursing homes may be dependent on prevalence of cases in the surrounding community and infection control opportunities.

## INTRODUCTION

Since the introduction of severe acute respiratory syndrome coronavirus 2 (SARS-CoV-2), many nursing homes (NHs) worldwide have been hit by outbreaks of this new virus. In the Netherlands, it is currently advised to perform SARS-CoV-2 testing in case of symptoms consistent with possible coronavirus disease 2019 (Covid-19). However, transmission of SARS-CoV-2 by a-and presymptomatic NH residents has been reported, which may warrant alternative screening policy in NHs.[1, 2] We aimed to assess the contribution of a- and presymptomatic residents and healthcare workers in transmission of SARS-CoV-2 in three NHs.

## METHODS

We conducted two serial point-prevalence surveys among residents and healthcare workers of three NHs in the Netherlands with recent SARS-CoV-2 introduction (i.e. at least one SARS-CoV-2 positive resident): NH-A, NH-B, and NH-C, of which NH-B had a Covid-19 outbreak in the month preceding the study and recent newly identified cases. NH characteristics including Covid-19 policy at the time of study conduction are presented in the *Table 1*.

**Table 1.** Characteristics of the participating NHs.

|  | **NH-A** | **NH-B** | **NH-C** |
| --- | --- | --- | --- |
| **Geographic characteristics** | Located in suburb of large city in the province of South Holland | Located in a village in the province of North Brabant | Located in suburb of large city in the province of North Holland |
| **Number of beds** | 140 | 120, and day service for people with dementia. | 121 |
| **Number and type of wards** | 8 wards (on 10 floors); 5 where psychogeriatric care is provided, 3 where somatic care is provided. | 15 residential groups with a separate front door each; 12 where psychogeriatric care is provided, 2 where somatic care is provided, 1 where both psychogeriatric and somatic care are provided. | 14 wards (on 5 floors); 3 where psychogeriatric care is provided, 4 where somatic care is provided, 2 where psychiatric care is provided, 2 where geriatric rehabilitation is provided, 1 where long-term care for acquired brain injury is provided and 1 where intermediate care is provided. |
| **Covid-19 policy at the time of study conduction \*** | Preventive use of surgical masks by healthcare workers on all wards; use of isolation gown, gloves over the wrists, goggles and a face mask in contact with Covid-19 suspected or confirmed residents. Covid-19 confirmed residents are transferred to a designated Covid-19 cohort ward; residents on the former ward of Covid-19 confirmed residents are placed in quarantine for 14 days. | Residential groups are divided in two cohorts because of previous Covid-19 cases; one cohort without and one cohort with Covid-19 (suspected) residents. Covid-19 suspected or confirmed residents are isolated in their rooms (or a complete residential group is isolated in case of multiple cases); use of isolation gown, gloves over the wrists, goggles and a face mask in contact with these residents. | Preventive use of face masks and gloves at the intermediate care ward. Covid-19 confirmed residents are isolated in their rooms (or a complete ward is isolated in case of multiple cases); use of isolation gown, gloves over the wrists, goggles and a face mask in contact with these residents. On one ward reversed isolation (negative residents isolated in their rooms) because of the inability to instruct a positive resident. |

NH= nursing home, PPE= personal protective equipment, Covid-19= Coronavirus disease 2019.
\*With regard to healthcare workers, policy of the National Institute for Public Health and the Environment is followed, implying that all individuals with Covid-19 consistent symptoms stay home until they receive their SARS-CoV-2 test results. In case of a negative result, healthcare workers with mild symptoms are allowed to work, taking into account general hygiene precautions. Health care workers with a positive test stay home until minimal 7 days after symptom onset, and until they have been free of fever for at least 48 hours and free of symptoms for at least 24 hours. In exceptional situations, positive and symptomatic healthcare workers can work using PPE. In addition to the abovementioned policy, policy of the participating NHs in case of positive but asymptomatic healthcare workers was that these individuals stay home for 72 hours; national policy as stated above was followed if symptoms developed in the meantime, if no symptoms developed healthcare workers were allowed to work using PPE.

Point-prevalence surveys included SARS-CoV-2 testing of all residents and healthcare workers, irrespective of whether they had been previously tested SARS-CoV-2 positive, and questionnaire completion. The first survey was performed in the week of May 4th, 2020, and was repeated 7 days later when tests were negative. SARS-CoV-2 positive individuals without symptoms or with atypical symptoms were followed 14 days for development of symptoms.

Nasopharyngeal and oropharyngeal swabs were collected by trained healthcare workers and specialized swab teams from the Public Health Service, in accordance with national guidelines.[3] Samples were transported to collaborating laboratories at the end of each test day, where they were tested for SARS-CoV-2 polymerase chain reaction (PCR) targets, see *Supplementary Material*.

Questionnaires were completed on the day of SARS-CoV-2 testing. Resident questionnaires were completed by healthcare workers, based on an interview with the resident and/or review of medical records; healthcare worker questionnaires were completed by the research team based on an interview with the healthcare worker, or online by healthcare workers themselves. The questionnaire included a standardized symptom-assessment form (for included signs and symptoms, see below). For residents, additionally documented were: ward name and type, previous Covid-19 disease, and recent admission or internal relocation. Healthcare workers were additionally asked on their profession, wards where they had worked the preceding 14 days, and personal protective equipment (PPE) use.

A participant was classified symptomatic in case of at least one new/worsened typical or atypical symptom of Covid-19 in the 14 days before a positive SARS-CoV-2 test. Typical symptoms included fever (T>38.0°C (100.4°F)), cough, and shortness of breath. Atypical symptoms included chills, malaise, fatigue, rhinorrhea, nasal congestion, sore throat, myalgia, headache, nausea or diarrhea, diminished intake, and loss of smell or taste. For residents, (increased) confusion and decreased saturation were also classified as atypical. A participant was classified presymptomatic if no symptoms were present the 14 days before a positive SARS-CoV-2 test, but typical or atypical symptoms developed during follow-up; when no symptoms developed during follow-up the participant was classified asymptomatic.

SARS-CoV-2 testing was in accordance with local policy of the three NHs. Residents, or their representatives in case of legal incapacity, were given the opportunity to opt-out for using their data in the study. Health care workers were asked informed consent for using their data prior to questionnaire administration. The Medical Ethics Committee of the VU University Medical Centre in of the Medical Research Involving Human Subjects Act.

## RESULTS

A total of 297 NH residents were included in the study (overall response: 86%; NH-A: 86%, NH-B: 90%, NH-C: 80%). 542 healthcare workers were included (overall response: 91%; NH-A: 94%, NH-B: 93%, NH-C: 87%). Demographic characteristics are presented in *Table 2 and 3*.

**Table 2.** Demographic characteristics of residents of participating NHs.

|  | **NH-A (N=114)** | **NH-B (N=104)** | **NH-C (N=79)** |
| --- | --- | --- | --- |
| **Age, average (range)** | 86.2 (68 – 103) | 84.9 (55 – 103) | 79.5 (47 – 102) |
| **Female, n (%)** | 84 (73.7) | 84 (80.8) | 51 (64.6) |
| **Care type\* :** |  |  |  |
| Geriatric rehabilitation / short term care, n (%) | N/A | N/A | 20 (24.1) |
| Long-term somatic care, n (%) | 37 (32.5) | 18 (19.6) | 28 (35.4) |
| Long-term psychogeriatric care, n (%) | 77 (67.5) | 74 (80.4) | 17 (21.5) |
| Psychiatric care, n (%) | N/A | N/A | 9 (11.4) |
| Intermediate care, n (%) | N/A | N/A | 5 (6.3) |
| **Previous or current Covid-19 (%)** | 0 | 13 (12.5) | 3 (3.8) |
| Covid-19 based hospitalization, n (%) | 0 | 0 | 0 |
| **Recent (<14 days) admission from other healthcare institution, n (%)** | 0 | 2 (1.9) | 11 (13.9) |
| **Recent (<14 days) internal relocation, n (%)** | 1 (0.9) | 1 (1.0) | 9 (11.4) |
| **Comorbidity:** |  |  |  |
| Pulmonary disease, n(%) | 19 (16.7) | 12 (11.5) | 10 (12.7) |
| Cardiovascular disease, n (%) | 51 (44.7) | 28 (26.9) | 18 (22.8) |
| Cerebrovascular disease, n (%) | 27 (23.7) | 23 (22.1) | 18 (22.8) |
| Diabetes, n (%) | 28 (24.6) | 17 (16.3) | 27 (34.2) |
| Dementia / cognitive impairment, n (%) | 85 (74.6) | 83 (79.8) | 24 (30.4) |
| Reduced kidney function, n (%) | 15 (13.2) | 4 (3.8) | 11 (13.8) |
| Obesity, n (%) | 11 (9.6) | 4 (3.8) | 12 (15.2) |

\*In NH-B, data on care type was missing for 12 residents.

**Table 3.** Demographic characteristics of healthcare workers of participating NHs.

|  | **NH-A (n=185)** | **NH-B (n=184)** | **NH-C (n=177)** |
| --- | --- | --- | --- |
| **Age, average (range)** | 46.7 (18 – 68) | 43.0 (18 – 67) | 47.5 (18 – 71) |
| **Female, n (%)** | 175 (94.6) | 167 (90.8) | 153 (86.4) |
| **Profession<sup>\*</sup>:** |  |  |  |
| Nurse aid /nurse assistant, n (%) | 90 (48.7) | 94 (51.1) | 63 (35.6) |
| Nurse, n (%) | 17 (9.1) | 24 (13.0) | 33 (18.6) |
| Physical therapist, n (%) | 2 (1.1) | 3 (1.6) | 6 (3.4) |
| Physician, n (%) | 3 (1.6) | 3 (1.6) | 7 (4.0) |
| Other, n (%) | 73 (39.5) | 60 (32.6) | 66 (37.3) |
| **PPE use<sup>\*\*</sup>:** | (N=69) | (N=61) | (N=61) |
| Face mask, n (%) | 68 (98.6) | 53 (86.9) | 49 (80.3) |
| Goggles, n (%) | 33 (47.8) | 51 (83.6) | 41 (67.2) |
| Gloves, n (%) | 67 (97.1) | 53 (86.9) | 54 (88.5) |
| Isolation gown, n (%) | 59 (85.5) | 50 (82.0) | 51 (83.6) |
| None, n (%) | 1 (1.4) | 7 (11.5) | 7 (11.5) |

NH= nursing home, Covid-19= Coronavirus disease 2019, PPE= personal protective equipment.
<sup>\*</sup>In NH-C, data on profession was missing for 2 healthcare workers.
<sup>\*\*</sup>In nurse aids / nurse assistants and nurses who indicated to have worked on wards with Covid-19 suspected or confirmed residents.

At the first point-prevalence survey, 15 residents (5%) tested SARS-CoV-2 positive (NH-A: 1, NH-B: 11, NH-C: 3), of which 9 (NH-B: 7, NH-C: 2) had a previous positive test result before study onset. Of the 6 newly identified cases of SARS-CoV-2, 2 were symptomatic (both in NH-B; Cycle threshold (Ct) value 22 in both), 1 was presymptomatic (in NH-A; Ct value >35), and 3 remained asymptomatic during follow-up (2 in NH-B and one in NH-C; Ct values 23, 30, and 32). A total of 8 healthcare workers tested SARS-CoV-2 positive at the first point-prevalence survey (NH-B: 7, NH-C: 1). All had typical symptoms; 5 had not worked in the two weeks before the first point-prevalence survey. At the second point-prevalence survey, one resident and one healthcare worker in NH-A tested SARS-CoV-2 positive (Ct value >35 and 24, respectively), both were asymptomatic on the day of testing.

## DISCUSSION

We aimed to study transmission of SARS-CoV-2 in three NHs with recent introduction of the virus, and to determine the role of a- and presymptomatic residents and healthcare workers herein. We identified 3 asymptomatic and 1 presymptomatic SARS-CoV-2 positive residents at the first point-prevalence survey. However, the number of cases identified in the follow-up survey was low (i.e. 2 in NH-A, none in NH-B and NH-C), which indicates very limited transmission and impeded the ability to answer the aforementioned research question. Nevertheless, we identified an asymptomatic resident and healthcare worker with Ct values below 25, suggesting that these cases have the potential to contribute to viral spread as previously suggested. [1, 2, 4]

Interestingly, the introduction of SARS-CoV-2 in two of the participating NHs (i.e. NH-A and NH-C) has not resulted in facility wide outbreaks during the study and the weeks thereafter. NH-B had an outbreak before the start of the study, and newly identified cases shortly before study onset. In response to the previous outbreak, this NH had taken measures such as increased hygiene precautions, the setup of cohorts of SARS-CoV-2 positive residents, and screening of healthcare workers for SARS-CoV-2, regardless of presence of symptoms (see also *Table 1*). The current study suggests that these measures were effective in this NH since no new transmission had occurred the weeks after the identification of new cases.

Other factors that may have contributed to the prevention of spread in the participating NHs include the decreased prevalence of Covid-19 in the Netherlands since mid-April,[5] reducing chances of new introductions of the virus in NHs. In addition, whereas NHs were – due to scarcity of Covid-19 tests – previously advised to stop performing Covid-19 tests after two positive cases (and consider all symptomatic residents in that ward Covid-19 positive), the availability of Covid-19 tests has increased since April 10th. NHs have been able to perform low threshold testing ever since, facilitating early recognition of cases and, if appropriate measures are taken in response, decreasing chances of viral spread. Likewise, previously scarcely available PPE have become available on larger scale for NHs since April 13^th^. We indeed found high PPE use in the participating NHs (*Table 3*) and although we did not evaluate whether they were appropriately used, it is plausible that this contributed to the prevention of further spread. Finally, the constructional features of the NHs may have been beneficial in preventing transmission, e.g. by the ability to physically separate wards, and an interior that facilitates quarantine and isolation measures. We did not collect data on the quality of the buildings’ ventilation systems; it may be interesting to include this in future studies, given previous calls to include building engineering controls as part of the infection control strategy.[6]

Our study, *Roxby et al.[7]* reported limited detection and transmission of SARS-CoV-2 in a NH with recent Covid-19 cases, upon screening of all residents and healthcare workers.[7] There were asymptomatic residents among the identified cases, like in our study, which one may argue calls for policy beyond symptom-based screening. This is in line with recommendations of the European Center for Disease Prevention and Control (ECDC) to test all residents and healthcare workers once a confirmed case is detected (and in areas with ongoing community transmission, to test healthcare workers regularly even without any confirmed cases).[8] On the other hand, authors of a French study in which all healthcare workers of a NH were tested for SARS-CoV-2 after a first positive resident, argued that human and financial resources for systematic screening are disproportionate to its effectiveness (they identified only one asymptomatic individual).[9]

## CONCLUSIONS AND IMPLICATIONS

Based on our findings, we argue that the necessity of large-scale screening in NHs may be dependent of the local situation regarding prevalence of cases in the surrounding community and infection control opportunities. If availability of equipment and constructional features facilitate rapid application of appropriate measures after a first identified case, this may suffice in preventing further transmission of SARS-CoV-2.

In conclusion; although we were not able to answer our predefined research question on the contribution of a-and presymptomatic cases in transmission of SARS-CoV-2, our study confirms a-and presymptomatic occurrence of Covid-19 among residents and healthcare workers. We described factors that may contribute to the prevention of SARS-CoV-2 transmission in NHs. Finally, our findings add to the discussion of effective Covid-19 screening policy in NHs.

## Data Availability

The datasets generated during and/or analysed during the current study are available from the corresponding author on request

## Acknowledgements

We thank all nurses and physician assistants of the participating NHs and Public Health Services for their contribution to and/or performance of SARS-CoV-2 testing. We thank the persons from Amsterdam UMC who assisted in questionnaire administration. We thank the NHs for their participation in the study in general, and in particular those individuals involved in logistic and/or administrative aspects of the study. We thank the laboratories of the Franciscus Gasthuis & Vlietland, Erasmus Medical Center, Public Health Service of Amsterdam, Wageningen Bioveterinary Research, and Microvida for the performance of SARS-CoV-2 tests.

## REFERENCES

1. Arons MM, Hatfield KM, Reddy SC, Kimball A, James A, Jacobs JR et al. Presymptomatic SARS-CoV-2 Infections and Transmission in a Skilled Nursing Facility. New England Journal of Medicine. 2020;382(22):2081-90. doi:10.1056/NEJMoa2008457.

2. Goldberg SA, Pu CT, Thompson RW, Mark E, Sequist TD, Grabowski DC. Asymptomatic Spread of COVID-19 in 97 Patients at a Skilled Nursing Facility. Journal of the American Medical Directors Association. 2020. doi:10.1016/j.jamda.2020.05.040.

3. Jacobi A, Meijer A, Molenaar P, van der Pol I. Afnametechniek specifieke viral diagnostiek (2019-nCoV, influenza)>. National Institute for Public Health and the Environment (RIVM).. https://lci.rivm.nl/sites/default/files/2020-02/Influenza%20%26%20COVID-19%20Afnametechniek%20specifieke%20virale%20diagnostiek.pdf. Accessed 17 June 2020.

4. Bullard J, Dust K, Funk D, Strong JE, Alexander D, Garnett L et al. Predicting infectious SARS-CoV-2 from diagnostic samples. Clinical infectious diseases: an official publication of the Infectious Diseases Society of America. 2020. doi:10.1093/cid/ciaa638.

5. (RIVM). NIfPHatE. COVID-19 in graphs. https://www.rivm.nl/coronavirus-covid-19/grafieken. Accessed 15 June 2020.

6. Morawska L, Tang JW, Bahnfleth W, Bluyssen PM, Boerstra A, Buonanno G et al. How can airborne transmission of COVID-19 indoors be minimised? Environment international. 2020;142:105832. doi:10.1016/j.envint.2020.105832.

7. Roxby AC, Greninger AL, Hatfield KM, Lynch JB, Dellit TH, James A et al. Outbreak Investigation of COVID-19 Among Residents and Staff of an Independent and Assisted Living Community for Older Adults in Seattle, Washington. JAMA Internal Medicine. 2020. doi:10.1001/jamainternmed.2020.2233.

8. Team EPHE, Danis K, Fonteneau L, Georges S, Daniau C, Bernard-Stoecklin S et al. High impact of COVID-19 in long-term care facilities, suggestion for monitoring in the EU/EEA, May 2020. Eurosurveillance. 2020;25(22):2000956. doi:doi:https://doi.org/10.2807/1560-7917.ES.2020.25.22.2000956.

9. Guery R, Delaye C, Brule N, Nael V, Castain L, Raffi F et al. Limited effectiveness of systematic screening by nasopharyngeal RT-PCR of medicalized nursing home staff after a first case of COVID-19 in a resident. Médecine et Maladies Infectieuses. 2020. doi:https://doi.org/10.1016/j.medmal.2020.04.020.

